# Epilepsy surgery in children with operculo-insular epilepsy: Results of a large unicentric cohort

**DOI:** 10.1101/2024.05.15.24307360

**Authors:** Kudr Martin, Janča Radek, Jahodová Alena, Bělohlávková Anežka, Ebel Matyáš, Maulisová Alice, Bukačová Kateřina, Tichý Michal, Libý Petr, Kynčl Martin, Holubová Zuzana, Šanda Jan, Ježdík Petr, Ramos Rivera Gonzalo Alonso, Kopač Luka, Kršek Pavel

## Abstract

**Objective:** Epilepsy surgery in the operculo-insular cortex is challenging due to the difficult delineation of the epileptogenic zone and the high risk of post-operative deficits following resections in this region.

**Methods:** Pre- and post-surgical data from 30 pediatric patients who underwent opercular-insular cortex surgery at Motol Epilepsy Center Prague from 2010 to 2022 were analyzed.

**Results:** Focal cortical dysplasia (FCD, n = 15) was the predominant cause of epilepsy in the patients studied, followed by epilepsy-associated tumors (n = 5) and tuberous sclerosis complex (n = 2). In eight patients where FCD was the most likely etiology, histology was negative. The epileptogenic zone was in the dominant hemisphere in 16 patients. Variability in seizure semiology and electrophysiological findings necessitated multimodal imaging and advanced post-processing for precise epileptogenic zone localization. Stereoelectroencephalography (SEEG) monitoring was used in 23 patients. The use of oblique electrodes as guides during resection proved beneficial for the neurosurgeon. At the two-year follow-up, 23 patients were seizure-free (ILAE Classification of Outcome 1), and seven experienced a seizure frequency reduction of > 50% (ILAE 4). Nineteen remained seizure-free following the definitive outcome assessment (2–13 years post-surgery). Five from 15 patients operated in posterior insula experienced pyramidal tract ischemia, leading to permanent mild hemiparesis in three patients and moderate hemiparesis in one patient. One patient experienced unexpected pontine ischemia 9 days post-surgery, leading to permanent spastic hemiparesis.

Five other patients experienced transient deficits.

**Significance:** Despite diagnostic and surgical challenges, epilepsy surgery in the opercular-insular cortex can lead to excellent patient outcomes. A comprehensive diagnostic approach is crucial for surgical success. For surgeries in the posterior insula, ischemia in the pyramidal tract and subsequent hemiparesis of varying severity should be anticipated. However, in pediatric patients, there is a great chance for significant recovery with proper rehabilitation.

**Key points:**

- Despite the diagnostic and surgical challenges, patients who underwent operculo-insular cortex surgery achieved excellent outcomes.
- Seizures from the operculo-insular region exhibit diverse semiology and propagation patterns (i.e., frontal, perisylvian, and temporal).
- An initially negative MRI finding is common. Video EEG typically reveals an extensive finding. A multimodal diagnostic approach is crucial.
- SEEG was necessary in a significant number of cases, and the use of oblique electrodes as guides proved beneficial for the neurosurgeon.
- For surgeries in the posterior insula, ischemia in the pyramidal tract and subsequent hemiparesis of varying severity should be anticipated.

## INTRODUCTION

The insula was likely first described by Vicq d’Azyr as “the convolutions situated between the Sylvian fissure and the corpus striatum”^1^. In 1809, the German physician and anatomist Dr. Johann Christian Reil introduced the concept of the insular lobe, which was subsequently named the “Island of Reil” in the first edition of Gray’s Anatomy in 1858^2^. In 1896, Tracy Earl Clark was the first to describe the insula as “the fifth lobe of the brain”^3^.

Due to insights gained from autopsies, the operculo-insular cortex anatomy has been understood since it was first researched. However, understanding of its function continues to evolve. Initial hypotheses in the second half of the 19th century discussed its connection to speech expression; however, these hypotheses were refuted by Broca. Contemporary knowledge states that the operculo-insular cortex is significantly connected with other brain lobes, playing an integrative role^4^. It links information from diverse functional systems, including the social-emotional, sensorimotor, olfacto-gustatory, and cognitive domains. This role was identified following a meta-analysis of 1,786 functional neuroimaging experiments^5^.

The concept of insular epilepsy was first discussed by Guillaume and Mazaras in the late 1940s, followed by Penfield, Jasper, and Faulk in the 1950s^6–9^. The concept stemmed from the analysis of electrocorticography (ECoG) records of patients with temporal lobe epilepsy coupled with observations of ictal symptoms akin to spontaneous seizures during direct electrical stimulation of the insular cortex. However, despite systematic efforts, spontaneous epileptic discharges originating focally in the insular cortex^10^ were never recorded by Penfield et al. In the subsequent decades, limited case reports of epilepsy associated with the operculo-insular cortex were published. The pioneering work of Isnard et al. demonstrated the feasibility of stereoelectroencephalography (SEEG) exploration of the operculo-insular cortex^11^. The researchers also described a stereotyped sequence of ictal symptoms associated with intrainsular discharges, which included laryngeal constriction, unpleasant perioral paresthesia, lateralized somatosensory sensations, dysarthria, and focal somatomotor signs^10^. Subsequent studies have identified additional ictal symptoms typical of insular seizures^12–15^. This accumulating evidence suggests that insular epilepsy is a significant mimicker, with its semiology dependent on the pattern of seizure spread. This conclusion was further validated in a comprehensive review by Jobst et al.^16^, which noted that besides the typical “perisylvian” clinical pattern, insular seizures can mimic temporal and frontal seizures and even epileptic spasms.

Delineating the epileptogenic zone (EZ) in the operculo-insular region is challenging during pre-surgical evaluation. SEEG is the method of choice for patients with operculo-insular epilepsy. Using a combination of orthogonal and oblique electrodes is particularly advantageous and has been proven safe^17^. Such exploration must be grounded in a robust semiological hypothesis regarding the seizure onset and its propagation, as well as on non-invasive diagnostic method results^16^.

Surgical resection is another challenging task associated with this region, as surgery in the operculo-insular region carries significant risks due to its proximity to the language processing cortex, the perisylvian blood vessels laterally, and the pyramidal tract and basal ganglia medially.

Published cohorts of pediatric patients who have undergone epilepsy surgery in the operculo-insular region are smaller than those of adult patients^18^. This is particularly interesting, considering that children represent an excellent group of candidates for epilepsy surgery, and positive outcomes can be anticipated. Additionally, due to the higher plasticity of the brain, children are less likely to experience permanent post-operative functional deficits.

This manuscript presents a large unicentric cohort of pediatric patients who underwent epilepsy surgery in the operculo-insular region.

## METHODS

### Patients

In a cohort of 341 pediatric patients who underwent resective epilepsy surgery at the Motol Epilepsy Center between 2010 and 2022, we retrospectively identified 30 patients who underwent operations in the opercular-insular region, either exclusively or as part of a larger resection. Patients who underwent larger disconnective procedures, such as hemispherectomy, were not included. Information regarding medical history, neurological status, and laterality was collected from hospital records. The study was approved by the institutional Ethical committee of Motol University Hospital (2022/06/15 - EK - 602.24/22).

### Semiology

Semiology was re-evaluated in every case according to the ILAE classification of seizure types^19^, utilizing medical history data and the results of video electroencephalography (VEEG). In each case, seizure details were further assessed. If a patient experienced multiple types of seizures, these were documented. For patients who underwent reoperation, we used the new semiology following the initial surgery.

A semiquantitative analysis of the presence of semiological signs was conducted for patients who underwent surgeries in the anterior, posterior, and junction of both regions, which represent anatomical insular subgroups. In addition to the different seizure types defined by the ILAE, we included semiological signs that we consider important in the context of operculo-insular epilepsy, namely epigastric aura, laryngeal constriction, non-specific aura, aphasia, and hemiparesis. In each subgroup defined by the surgery localization, the total frequency of semiological signs and their frequency as the initial sign were calculated.

Based on seizure semiology, one or more seizure propagation patterns were assigned to every case as previously described (i.e., frontal, perisylvian, temporal, and spasm pattern)^16^.

### VEEG

VEEG was conducted in all patients except for one case with a benign tumor that was operated on based on neuroimaging and a routine scalp EEG. The original VEEG descriptions and files for each patient were analyzed. Ictal and interictal EEG patterns were localized according to their onset in one or more of the five regions of both hemispheres. The localizations used were frontal, central, temporal, parietal, and occipital. Additionally, hemispheric, bilateral, and generalized findings were observed. For practical purposes, the VEEG findings were categorized into three broader groups: lobar, multilobar, and bilateral.

### MRI

High-resolution brain magnetic resonance imaging (MRI; 1.5 T and subsequently 3 T, <1 mm^3^ isotropic) with a dedicated epilepsy protocol was performed in all cases and reviewed by experienced neuroradiologists (MK and ZH). All MRI examinations were consistent with the recommendations of the ILAE regarding the use of MRI in the care of patients with epilepsy^20^.

### Functional neuroimaging methods

18F-Fluoro-deoxy-glucose positron emission tomography (FDG-PET) registered to MRI was realized to identify hypometabolic cortexes. Additionally, partial volume effect correction of PET (pvcPET) was performed to increase limited PET resolution and identify subtle lesions in recent cases after 2020^21–23^.

Subtraction ictal single photon emission computed tomography (SPECT; 99mTc-ECD and subsequently 99mTc-HMPAO) co-registered to MRI (SISCOM) was conducted to localize ictal hyperperfusion in selected cases. Diffusion tensor imaging (DTI) of the pyramidal tract and the arcuate fasciculus was routinely incorporated as part of the surgical neuronavigation process, as well as functional MRI for localizing the language and motor cortex.

### Neuropsychological assessments

Patients in our epilepsy surgery program were routinely neuropsychologically tested. The data used here are values obtained from several age-dependent tests: the Bayley Scale of Infant and Toddler Development^24^, the Wechsler Preschool and Primary Scale of Intelligence^25^, and the Wechsler Intelligence Scale for Children^26^. Neuropsychologists (AM and KB) routinely performed pre-surgery and 1-year post-surgery testing and retested individual cases at longer follow-ups.

### Intracranial EEG

Long-term intracranial video/EEG monitoring (iEEG) was indicated if (1) the patient had normal or non-localizing findings on brain MRI (e.g., a subtle/questionable lesion, a lesion with ill-defined borders, or multiple lesions), (2) the structural abnormality was adjacent to eloquent cortical areas, and (3) the findings of non-invasive examinations were discordant.

The implantation scheme of the depth (SEEG) electrodes (Dixi Medical) was planned using the neuronavigation system StealthStation S7 (Medtronic) for frame non-robot Cosman-Robert-Wells stereotaxy. Orthogonal and oblique trajectories were realized for dense insular exploration. Early cases underwent implantation with a combination of depth and subdural electrodes (Integra LifeSciences).

Computed tomography scans were performed to exclude complications and were registered to pre-operative MRI for the anatomical localization of all recording contacts immediately after implantation^27^. Recorded iEEGs (Stellate Harmony or Natus Quantum) lasted 6–14 days, depending on the habitual frequency of a patient’s seizures and the schedule of planned electrophysiological studies, such as extraoperative electrical stimulation mapping. Standard interictal and ictal iEEG assessment was extended using computerized quantitative analyses^28–30^.

### Surgery

The presumed localization of the EZ resulted from electrophysiological (iEEG and stimulation mapping) and neuroimaging (MRI, FDG-PET, and SISCOM) findings based on expert panel consensus; example case No. 9 is shown in Figure 1. The exact extent of the resection (presumed EZ) was marked in the neuronavigation system together with the eloquent cortex and DTI tracts.

**Figure 1.**
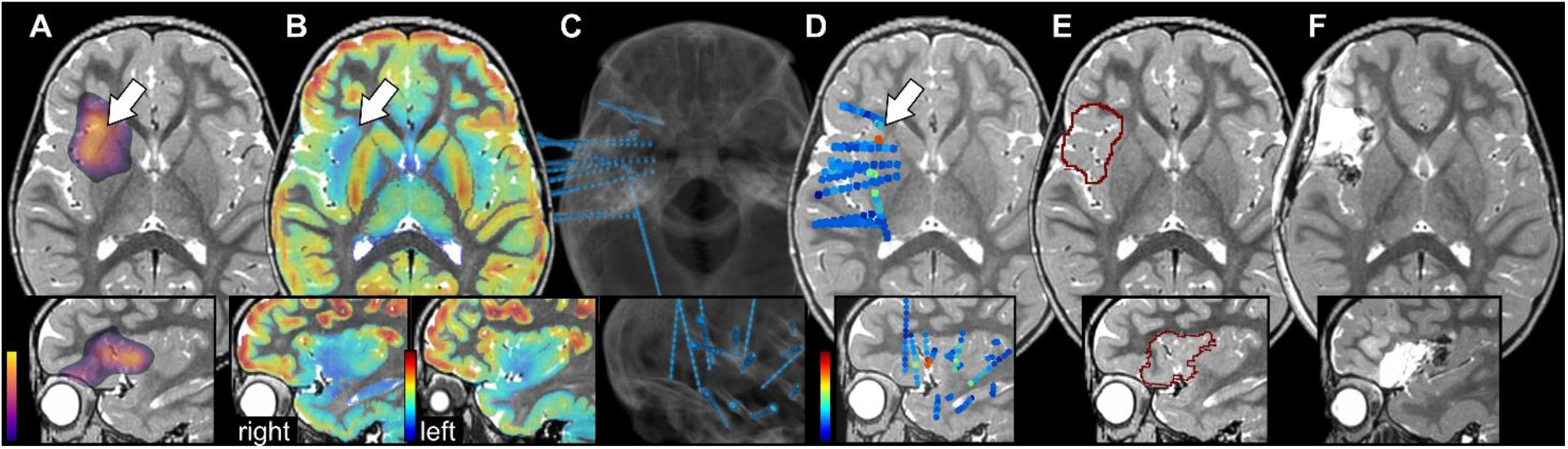
An example of the importance of multimodal assessment is seen in case No. 9, who presented with normal MRI findings. (A) SISCOM indicated ictal hyperperfusion (arrow) on the right anterior operculo-insular region. The axial view is supplemented by the sagittal view of the insula below. (B) FDG-PET with pvcPET post-processing showed hypometabolism (cold colors) in the same region (arrow). The comparison of the right and left insulas in the sagittal view helped to identify hypometabolic margins. (C) A post-implantation CT showed dense insular SEEG coverage using orthogonal and oblique trajectories (blue). (D) Visualization of the seizure onset zone (warm colors) resulted from computerized quantitative analysis. (E) The extent of the planned resection resulted from neuroimaging and electrophysiological examination. (F) Post-resection MRI. Note: All modalities were pre-processed for the 3D Slicer viewer.

In our center, surgical resection is performed immediately after the completion of invasive electrophysiological studies. This enables the preservation of SEEG electrodes as an intraoperative neurosurgery guide for resection in the anatomically complex opercular-insular region (see Figure 2)^31^.

**Figure 2.**
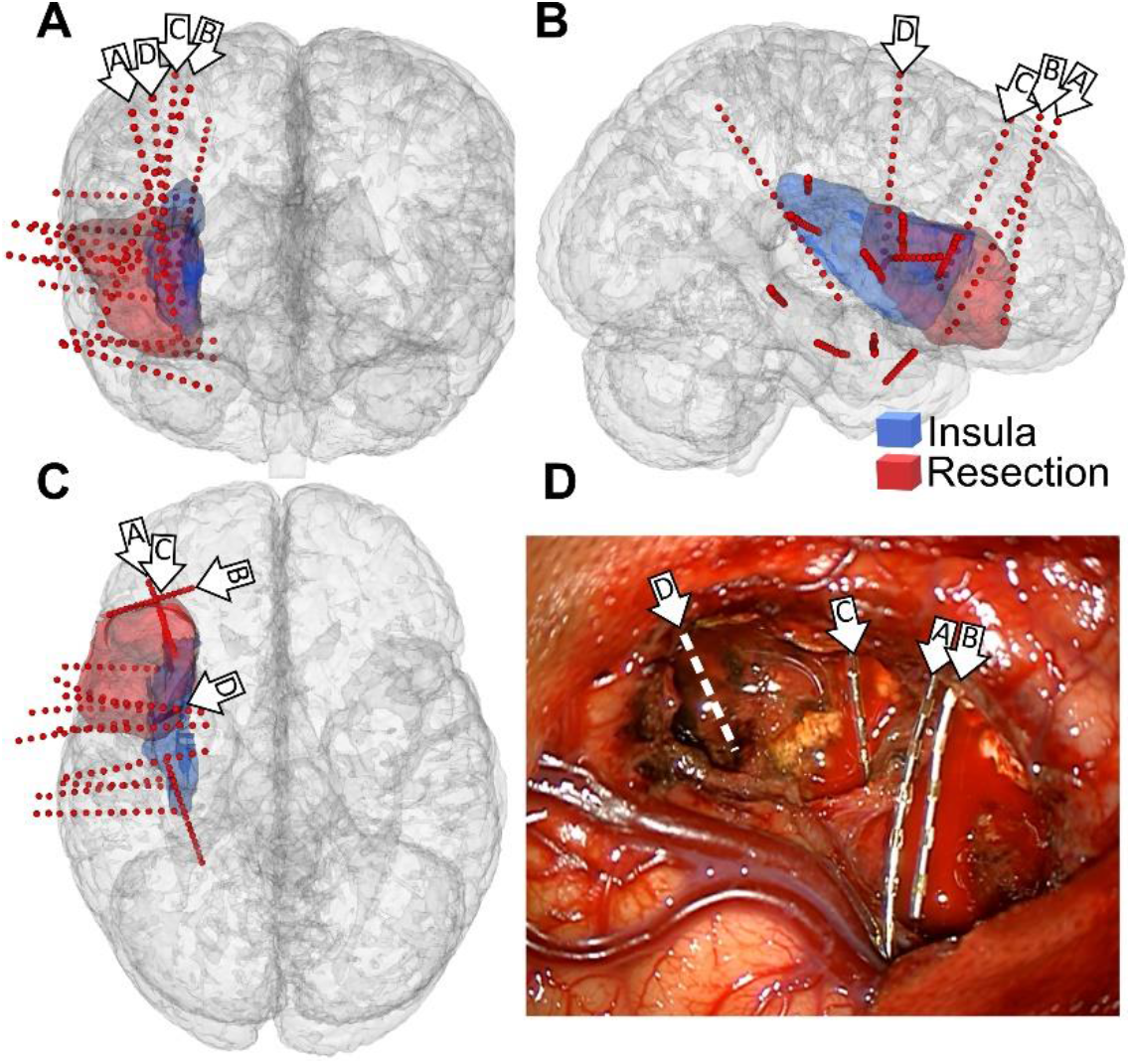
The benefit of preserving SEEG electrodes as a surgical guide to determine resection and anatomical margins is seen in case No. 9. (A–C) Coronal, sagittal, and axial views on 3D brain visualization. Red dots are the recording contacts of SEEG electrodes. The right insula is semitransparently marked by blue, and the resection area by red. Arrows with letters mark oblique electrodes preserved as surgical guides. (D) Photo taken with a neurosurgical microscope at the end of resection. Note: electrode “D” was explanted during the operation.

Continuous intraoperative monitoring of motor function was conducted in all patients due to the proximity of the primary motor area and the corticospinal tract, adhering to the electrical stimulation mapping (ESM) paradigm developed for the youngest patients^32^. If necessary, the procedure was combined with awake craniotomy due to the proximity of the EZ and eloquent speech centers.

A post-operative brain MRI was performed on all patients to assess the completeness of the resection and to exclude complications on the first post-operative day. Resections were considered complete if the region of significant intracranial EEG abnormality and MRI lesion (if present) was entirely removed^33^.

### Histology

Histological findings were originally evaluated according to Palmini’s classification of focal cortical dysplasias (FCDs) and were re-evaluated according to the ILAE classification used in this study^34,35^.

### Post-operative outcomes

Seizure outcomes were evaluated according to the ILAE classification 2 years post-surgery^36^. To provide the most relevant and clinically applicable information, the definitive outcome was assessed in all cases as of April 1, 2024. Post-operative deficit assessments were based on information obtained from regular post-operative neurological follow-ups, which were conducted at least every 4 months. Neuropsychological outcomes were compared between pre- and post-surgery testing at the 1-year follow-up.

## RESULTS

### Patients

Thirty patients were retrospectively enrolled in the study, comprising 16 males and 14 females. The left hemisphere was affected in 18 patients (60%) and the right in 12 (40%). Seizure onset in the dominant hemisphere was confirmed in 16 (53.3%) individuals. Seizures began at 4 years and 3 months old on average (range = 21 days to 16 years and 8 months). The average age at the time of the last surgery was 10 years and 11 months (range = 1 year and 5 months to 18 years and 7 months). Twenty (66.7%) patients exhibited normal neurological findings upon evaluation, while two patients (6.7%) had mild hemiparesis. A variety of less severe neurological symptoms were observed in eight (26.7%) cases (Supplementary Table S1).

The cohort was divided into cases involving operations at the anterior and posterior operculo-insular regions and at the border of these areas. The anterior operculo-insular region was defined as the anterior insula (short gyri), frontal operculum, and anterior part of the temporal operculum. The posterior region was defined by the posterior insula (long gyri), rolandic and parietal opercula, and the posterior part of the temporal operculum. Detailed information for each patient is provided in Table S1.

### Seizure semiology

The most frequent seizure types were tonic, automatism, sensory, autonomic, behavior arrest, hyperkinetic, and laryngeal constriction. Sensory seizures were the most common initial type, particularly in patients surgically who underwent surgery in the posterior part of the operculo-insular region. Laryngeal constriction was more prevalent in cases involving the posterior region, whereas epigastric aura, which was less frequently observed overall, was exclusively seen in patients who underwent anterior region surgery.

The most common seizure propagation pattern was frontal, followed by perisylvian and temporal patterns. Epileptic spasms occurred least often. When comparing individual groups, a higher representation of the perisylvian pattern and a lower representation of the temporal pattern was evident in patients who underwent posterior region surgery. Spasms were more commonly observed in cases operated on anteriorly than those operated on posteriorly. Details are summarized in Table 1 and Supplementary Tables S1 and S2.

**Table 1.** Summary of initial seizure semiology and propagation patterns observed in patients who underwent surgery on different operculo-insular regions.

| Operculo-insular region: | Ant. | A-P | Post. |
| --- | --- | --- | --- |
| Number of patients | 11 | 4 | 15 |
| **Initial semiology** |  |  |  |
| Focal motor |  |  |  |
| Automatisms | 4 | 0 | 2 |
| Atonic | 1 | 0 | 0 |
| Clonic | 1 | 0 | 2 |
| Epileptic spasms | 3* | 0 | 1 |
| Hyperkinetic | 2 | 0 | 1 |
| Myoclonic | 0 | 0 | 1 |
| Tonic | 3 | 2 | 5 |
| Focal nonmotor |  |  |  |
| Autonomic | 3 | 1 | 2 |
| Behavior arrest | 7 | 1 | 3 |
| Cognitive | 1 | 2 | 0 |
| Emotional | 3 | 0 | 3 |
| Sensory | 1 | 2 | 14* |
| Other |  |  |  |
| FBTCS | 0 | 0 | 0 |
| Epigastric aura | 3* | 0 | 0 |
| Laryngeal constriction | 1 | 2 | 3* |
| Non-specific aura | 0 | 0 | 2 |
| Aphasia | 0 | 0 | 2 |
| Hemiparesis | 0 | 0 | 1 |
| **Propagation pattern** |  |  |  |
| Frontal | 8 | 2 | 11 |
| Temporal | 7 | 3 | 5 |
| Perisylvian | 3 | 2 | 13* |
| Spasm | 4* | 0 | 1 |

Key: Ant. – anterior region; A-P – junction between anterior and posterior region; post. – posterior region; FBTCS – focal to bilateral tonic-clonic seizure (\*) indicates an at least three times more frequent occurrence of symptoms in the anterior region compared to the posterior region.

### VEEG

Findings from scalp VEEGs (29/30) were extensive in most cases. Regarding interictal findings, most were assessed as either multilobar (16 cases; 55.2%) or bilateral (seven cases; 24.1%), with six (20.7%) assessed as lobar findings. The pattern was similar for ictal findings. Nineteen (65.5%) were multilobar, four (13.8%) were bilateral, and six (20.7%) were lobar (Table 2).

**Table 2.** Summary of scalp video-EEG and neuroimaging findings.

| VEEG |  | n = 29 |
| --- | --- | --- |
|  | Ictal | Interictal |
| Lobar | 6 | 6 |
| Multilobar | 16 | 19 |
| Bilateral | 7 | 4 |
| MRI |  | n = 30 |
| FCD oper-insular | 14 |  |
| FCD temporal | 1 |  |
| Mild HS | 1 |  |
| Tumor | 5 |  |
| TSC | 2 |  |
| Normal | 7 |  |
| FDG-PET |  | n = 28 |
| Correct loc. | 16 |  |
| Correct lat. | 25 |  |
| Incorrect lat. | 1 |  |
| Normal | 2 |  |
| SISCOM |  | n = 17 |
| Correct loc. | 12 |  |
| Correct lat. | 1 |  |
| Incorrect lat. | 0 |  |
| Non-localizing | 4 |  |

Key: lat. – lateralization, loc. – localization, HS – hippocampal sclerosis, TSC – tuberous sclerosis complex; n – number of patients.

**Table 3.** Post-surgery histology, seizure outcome, and anti-seizure medication characteristics.

| **Histology** | **Negative** | **FCD type 1** | **FCD type 2** | **Tumors** | **TSC** |
| --- | --- | --- | --- | --- | --- |
|  | 8 | 5 | 10 | 5 | 2 |
|  | **Class 1** | **Class 2** | **Class 3** | **Class 4** | **Class 5** |
| **Seizure outcome**<br>**ILAE classification** | **Seizure**<br>**free** | **Only auras** | **1–3 sz. per**<br>**y.** | **4 sz. per y.**<br>**to 50%**<br>**reduction** | **50% reduction**<br>**to 100%**<br>**increase** |
| 2 years follow-up (n = 30) | 23 | - | - | 7 | - |
| 2–5 years follow-up (n = 9) | 7 | - | 1 | 1 | - |
| > 5 years follow-up (n = 21) | 13 | - | - | 7 | 1 |
| **ASM reduction** | **Withdrawn** | **Reduction** | **No**<br>**reduction** | **Re-**<br>**administered** |  |
| 2 years (n = 30) | 2 | 14 | 14 | - |  |
| 2–5 years (n = 9) | - | 4 | 5 | - |  |
| 5–11 years (n = 21) | 7 | 5 | 7 | 2 |  |

Key: FCD – focal cortical dysplasia; TSC – tuberous sclerosis complex; ASM – anti-seizure medication; sz. – seizure; y. – year.; n – number of patients.

### MRI

Seven patients (23.3%) displayed normal MRI results, while 23 (76.7%) had visible MRI abnormalities. The predominant abnormality detected was suspected FCD in the operculo-insular region, which was observed in 14 patients (46.7%). Additionally, dysplastic changes were identified in the temporal lobe in one case (3.3%). There was a single instance (3.3%) of mild hippocampal sclerosis and two patients (6.7%) of tuberous sclerosis complex (TSC). Neoplastic lesions were apparent in five patients (16.7%; Table 2).

### Functional neuroimaging methods

FDG-PET was performed in 28 patients (93.3%), revealing operculo-insular hypometabolism in 16 cases (57.1%) and correct lateralization in 25 (89.3%). One case (3.6%) showed incorrect lateralization. SISCOM was conducted in 17 (56.7%) patients, with operculo-insular hyperperfusion identified in 12 (70.6%) and correct lateralization in 13 (76.5%). Two patients (7.1%) exhibited normal FDG-PET results, while SISCOM was non-localized in 4 (23.5%; Table 2).

### SEEG, surgery, and post-operative complications

Seven patients (23.3%) underwent a one-stage resection, while 23 (76.7%) required long-term iEEG monitoring (21 by SEEG only and two by SEEG combined with subdural electrodes). The number of implanted electrodes increased with epilepsy surgery team experience from 2 to 18 electrodes (up to 8 in the opercular-insular cortex). No complications associated with iEEG monitoring were reported. Awake craniotomies were necessary in four cases (13.3%) for eloquent speech center mapping. Anterior insula region surgery was needed in 11 cases (36.7%), four (13.3%) required surgery at the junction of the anterior and posterior insula, and 15 cases (50%) underwent surgery in the posterior insula region. In 12 cases (40%), the extent of surgery exceeded the boundaries of the operculo-insular region, with the frontal lobe partially involved in six cases, the temporal lobe in five, and the parietal lobe in one. Further details are provided in Supplementary Table 1. The surgical approach to the insular cortex involved creating “windows” between the vessels on the opercular cortex, as shown in Figure 3.

**Figure 3.**
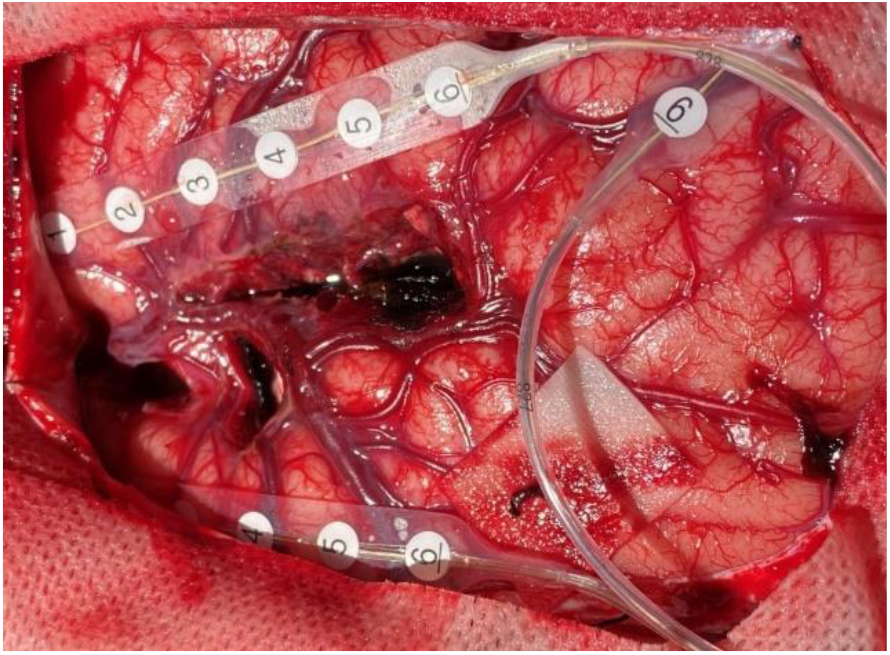
The surgical approach for the insular cortex involves creating “windows” between the vessels on the opercular cortex, as seen in case No. 30. The strips of electrodes used for post-resection electrocorticography (above the suprasylvian and infrasylvian cortex) and for intraoperative monitoring of motor functions (top right; over the primary motor cortex) are visible.

Post-operative complications were observed in nine cases (30%); four (13.3%) were temporary complications and five (16.7%) were permanent. The higher severity of deficiencies was related to the posterior operculo-insular surgery (four permanent and one temporary) compared to surgery in the anterior operaculo-insular region (one permanent and three temporary). Regarding anterior operculo-insular complications, one patient experienced a severe unexpected complication (pontine ischemia) nine days post-surgery in the form, leading to permanent spastic left-sided hemiparesis. Another patient had post-operative brain edema, which required a craniectomy and led to no permanent deficits. Of the patients who underwent posterior operculo-insular region surgery, five experienced ischemia in the pyramidal tract, resulting in hemiparesis. One of these patients recovered fully, one had permanent moderate hemiparesis (particularly distal on the right upper limb), and three had very mild hemiparesis after intensive rehabilitation. Details about the transient complications are provided in Supplementary Table S1.

### Histology

Histological analysis revealed FCD type 1 in five patients (16.7%) and FCD type 2 in 10 patients (33.3%). In eight patients (26.7%) with negative histopathological findings, FCD was suspected based on the combination of clinical features, neuroimaging results (3/8 structural and 7/8 functional), and intracranial EEG findings. Additionally, LEATs and TSC were identified in five (16.7%) and two patients (6.6%), respectively.

### Post-operative outcomes

Twenty-three patients (76.7%) were seizure-free at the 2-year follow-up (ILAE class 1). Seven patients (23.3%) demonstrated up to 50% reductions in seizure frequency (ILAE class 4). Regarding treatment adjustments, 14 patients (46.7%) at the 2-year follow-up underwent anti-seizure medication (ASM) tapering, while treatment was completely withdrawn in two patients (6.7%).

We evaluated definitive seizure outcomes after at least 5 years of follow-up. This outcome was achieved in 21 patients (5–13 years of follow-up): 13 patients (61.9%) were seizure-free, seven (33.3%) were ILAE class 4, and one (4.8%) was ILAE class 5. ASMs were completely withdrawn in seven patients (33.3%) and reduced in five (23.8%).

In the sub-group of 23 patients who underwent long-term iEEG monitoring, 14 (60.9%) were seizure-free upon evaluation of the definitive outcome. In these patients, we assumed a complete resection of the epileptogenic zone was achieved during the resection. For the remaining nine patients (39.1%) who experienced poorer outcomes, we considered the resection incomplete in five cases (55.6%) and complete in four (44.4%).

### Neuropsychological assessments

Of the 24 individuals who had accessible pre- and post-surgical neuropsychological data, six (25%) exhibited an over 10-point decrease in IQ 1-year post-operation. In two of these cases, the decline in overall IQ was associated with post-operative seizure occurrences. The remaining 18 (75%) patients either maintained or demonstrated improved IQ levels 1-year post-surgery.

## DISCUSSION

The complexity associated with epilepsy surgery in the operculo-insular area is due to the challenging diagnosis and the demanding nature of the surgical procedure itself, particularly for interventions in the posterior insula. These interventions are associated with a high risk of complications due to the risk of ischemia in the pyramidal tract. Although the number of studies focusing on epilepsy surgery in this region has increased in recent years, systematic assessments of pediatric patients remain scarce^37^. Our single-center study is one of the most extensive studies involving a pediatric cohort and long-term outcomes to date. Our results demonstrate that surgeries in this region can result in overall good outcomes with fewer permanent post-operative deficits than generally assumed.

From the perspective of EZ localization, the semiology of seizures, which is highly variable, can be misleading. This variability is influenced by two factors.

First, many functions (the social-emotional, sensorimotor, olfacto-gustatory, and cognitive cortex) are represented in a relatively small cortical area^5^. This corresponds with the wide variety of seizures described in our patient group. Across the group, sensory seizures of various types, behavior arrest, and tonic seizures were the most commonly reported initial symptoms. Automatisms, autonomic seizures, emotional seizures, and laryngeal constriction were also frequent, with other types of seizures occurring to a lesser extent. Regarding the difference between the anterior and posterior insula, a dominance of sensory seizures in epilepsy from the posterior operculo-insular region was noted, with laryngeal constriction being more common in posterior operculo-insular epilepsies or at their border. Epigastric aura was only found in patients who were operated on anteriorly.

Second, the insula is recognized as a “rich hub” that connects other parts of the brain^38^. Consequently, seizures originating in the insula can quickly spread to other areas. Jobst et al.^16^ identified four patterns of seizure spreading. In this study, the most commonly observed was the frontal spread pattern, followed by perisylvian and temporal patterns. Notably, in cases involving operations in the posterior operculo-insular region, the perisylvian pattern was dominant over the temporal pattern. The least common pattern were spasms, which occurred predominantly in cases involving anterior region operations compared to posterior cases, at a ratio of 4:1. It is also important to note the possible rapid spread to the contralateral insula, that has to be considered when planning SEEG exploration, monitoring, and subsequent surgery. This is demonstrated in the initially confusing MRI-negative patient (no. 15), where the semiology was non-lateralizing, scalp VEEG findings were bilateral or right-sided, FDG-PET showed right-sided hypometabolism, and the early iEEG resulted in additional SEEG electrode implantation to confirm seizure origin in the left hemisphere.

Given the insula’s location, standard scalp EEG and VEEG findings are typically extensive, with most findings being multilobar or bilateral^39^. Therefore, the MRI findings are crucial. The relatively high lesion detection rate in three-quarters of our patients^18^ was achieved by using a specialized MRI epilepsy protocol evaluated by experienced neuroradiologists. FCD was ascertained in two-thirds of these cases. However, the delineation of borders in these often subtle lesions was challenging.

The use of multimodal functional neuroimaging with advanced post-processing (pvcPET and SISCOM) proved beneficial, providing localization or lateralization in almost all cases where MRI results were normal. The role of functional imaging (functional MRI and DTI) in identifying eloquent areas (Broca’s and Wernicke’s areas, the pyramidal tract, and the arcuate fasciculus) was significant and helped with surgical planning. SEEG was necessary in three-quarters of cases and enabled the accurate localization and delineation of the EZ, with a clear trend towards more dense implanting of a greater number of electrodes over time.

Given the need to remove a narrow band of insular cortex with minimal damage to the medially located white matter, we consider the previously published method of leaving oblique electrodes as guides for the neurosurgeon during surgery to be crucial for surgical success^31^. This technique allows neurosurgeons to navigate much more precisely than with a standard neuronavigation system, mainly after brain tissue shift due to, e.g., cerebrospinal fluid leakage, edema, or anti-edema therapy. However, insular cortex resections remain one of the most challenging neurosurgical operations. In this surgery, neurosurgeons create “windows” between vessels over the opercular cortex.

FCD was the most frequent cause of epilepsy, being histologically confirmed in half of the patients. Although another tenth of the patients exhibited MRI signs of FCD (with varying reliability), their histology was normal. The high incidence of negative histological findings in our cohort might be due to insufficient tissue sampling because of cortex excavation using CUSA® (Integra LifeSciences). Additionally, the “discontinual” distribution of FCD in the cortex might contribute to the histological negativity.

Regarding post-operative outcomes, most patients were seizure-free. Two years post-surgery, 76.7% of patients were seizure-free, and another 23.3% reported at least a 50% reduction in seizure frequency. Although a slightly lower seizure-free rate (61.9%) was observed in the definitive outcomes subgroup (> 5 years follow-up), the benefit of epilepsy surgery was unequivocal. Although the insula is associated with various cognitive and emotional functions, our comparison of overall IQ pre- and 1-year post-surgery indicated stable or slightly improved performance in three-quarters of cases, with worsening only observed in cases with continued seizures. However, long-term cognitive outcomes must be comprehensively analyzed in another study.

One-third of our patients who underwent surgery in the posterior insula developed ischemia in the pyramidal tract, aligning with findings from previous studies^18^. Only one-fifth of patients experienced moderate hemiparesis, with the remainder presenting with either no or mild hemiparesis. This expected post-operative complication highlights the importance of detailed pre-operative counseling for parents and patients. It also emphasizes the need for well-planned post-operative rehabilitation strategies that can alleviate hemiparesis severity.

Surgical interventions in the operculo-insular region remain among the most formidable challenges in epilepsy surgery. Similarly, achieving a deeper understanding and conducting thorough investigations of the diverse functions of this fascinating brain region pose significant obstacles. We sincerely hope that our study will assist other epilepsy surgery teams in the diagnosis, planning, and performance of surgeries, ultimately leading to an improved quality of life for patients with epilepsy.

## CONCLUSION

Our large pediatric surgical study has helped to define phenotypes of children with opercular-insular epilepsy. The findings could be utilized to identify potential surgical candidates earlier. This early identification might be challenging as these patients typically experience seizures with diverse semiology, often presenting a wide array of semiological signs that include frontal (usually mesiofrontal), perisylvian, and temporal symptoms in addition to the typical insular symptoms. Moreover, these patients often exhibit initially negative MRI findings, and VEEG findings usually show extensive abnormalities.

Due to increasing knowledge, advances in electrophysiological and imaging methods, and neurosurgical techniques, these complex cases can now significantly benefit from timely epilepsy surgery. Our findings showed the benefit of using oblique SEEG electrodes as guides for neurosurgeons during surgical procedures. For surgeries in the posterior insula, ischemia in the pyramidal tract and subsequent hemiparesis of varying severity should be anticipated. These complications necessitate thorough pre-operative counseling for parents and patients and post-operative rehabilitation planning.

## Supporting information

Supplementry Table 1 (anonymized)

## Data Availability

Majority of data produced in the present work are contained in the manuscript and supplementary materials, potentially de-anonymizing information is available from the authors upon request

## Acknowledgments

This work was supported by the Ministry of Health of the Czech Republic NU23-08-00528; Ministry of Health of the Czech Republic, research project 00064203 Motol University Hospital; Charles University Grant Agency (GAUK), research project 666320. Radek Janca was supported by ERDF-Project Brain Dynamics, No. CZ.02.01.01/00/22_008/0004643. The authors thank CESNET for access to their data storage facility.

## Author Contributions

**Martin Kudr:** conceptualization (lead); writing – original draft (equal); writing – review & editing (equal); resources (equal). **Radek Janča:** conceptualization (supporting); writing – original draft (equal); writing – review & editing (equal); funding acquisition (equal); software (equal). **Alena Jahodová:** resources (equal). **Anežka Bělohlávková:** resources (equal). **Matyáš Ebel:** resources (equal); data curation (equal). **Alice Maulisová:** resources (equal). **Kateřina Bukačová:** resources (equal); writing – review & editing (supporting). **Michal Tichý:** resources (equal). **Petr Libý:** resources (equal). **Martin Kynčl:** resources (equal). **Zuzana Holubová:** resources (equal). **Jan Šanda:** resources (equal); data curation (equal). **Petr Ježdík:** software (equal); resources (equal); data curation (equal). **Rivera Gonzalo Alonso Ramos:** resources (equal). **Luka Kopač:** resources (equal). **Pavel Kršek:** conceptualization (supporting); supervision (lead); project administration (lead); funding acquisition (equal); writing – review & editing (equal); resources (equal).

## Conflicts of Interest

None of the authors has any conflict of interest to disclose.

## Ethical Publication Statement

We confirm that we have read the Journal’s position on issues involved in ethical publication and affirm that this report is consistent with those guidelines.

## Notes

### Competing Interest Statement

The authors have declared no competing interest.

### Author Declarations

The study was approved by the institutional Ethical committee of Motol University Hospital (2022/06/15 - EK - 602.24/22).

